# Mediation effects of inflammation on the association of physical activity and chronic kidney disease: evidence from questionnaire and device-measured assessments

**DOI:** 10.64898/2026.04.06.26350269

**Authors:** Xin Zhang, Ping Zeng

## Abstract

**Background:** The association between physical activity (PA) and chronic kidney disease (CKD) was initially explored; however, it remained unknown whether PA affected the incidence of CKD through the inflammation pathway. Moreover, objective PA measured by accelerometers was rarely considered for this association.

**Methods:** This study was performed in a large-scale prospective cohort with two different sub-cohorts: the International Physical Activity Questionnaire (IPAQ)-measured cohort (*N*=314,694); and the device-measured cohort (*N*=79,454). Cox models were conducted to assess the association of PA with incident CKD. The mediating role of inflammation in such association was investigated by four inflammation metrics [C-reactive protein (CRP), white blood cell (WBC), a low-grade inflammation (INFLA) score, and monocyte to high-density lipoprotein cholesterol ratio (MHR)].

**Results:** In the questionnaire-measured cohort, compared to low PA, moderate and high PA reduced the risk of CKD by approximately 28.0% (95% CI 24.4∼31.5%) or 37.6% (34.4∼40.7%), respectively. Inflammation significantly mediated this association, with the mediation proportion was 4.1% (3.0∼5.1%), 1.4% (1.1∼1.7%), 9.8% (7.7∼11.9%), and 1.4% (1.1∼1.7%) for CRP, WBC, INFLA score, and MHR, respectively. Evidence from the device-measured cohort further strengthened the robustness of our findings, but the effects were somewhat attenuated, with the mediation proportion being 2.2% (1.2∼3.2%), 0.8% (0.2∼1.3%), 4.3% (2.5∼6.0%), and 1.3% (0.6∼2.1%) for CRP, WBC, INFLA score, and MHR, respectively.

**Conclusions:** Our study reveals suggestive evidence for the association of active PA with reduced CKD risk and further demonstrates the mediating role of inflammation in such association, providing a novel perspective for the early prevention of CKD.

## Introduction

Over the past few years, chronic kidney disease (CKD) has rapidly become one of the global major contributors to years of life loss and mortality ^1^; ^2^. It is reported that approximately 850 million people around the world are affected by CKD, and the number is rising ^3^. Due to its irreversibility and slow progression ^4^, current treatments of CKD are mainly based on long-term conservative therapy ^2^, which imposes a huge economic burden on both patients and society ^1^. A complete understanding of risk factors related to CKD is essential to prevent this disease with effective interventions.

Physical activity (PA) is widely recognized as a key modifiable lifestyle factor that can help reduce the risk of CKD ^5–8^. Despite its importance, most existing studies have relied primarily on self-reported PA data collected through questionnaires. However, PA can be measured using two main methods—questionnaires and accelerometers—each with its distinct advantages ^7^; ^9^. Accelerometers offer a more precise quantification of PA duration and intensity, providing objective data that is particularly useful for examining dose-response relationships of different intensities of PA. Questionnaires excel at capturing detailed information on certain types of resistance exercise, which is more comprehensive. Therefore, combining PA data from both measurement methods allows for a more comprehensive understanding of the association of PA with CKD risk.

Additionally, although the potential mechanism of how PA influences the incidence of CKD is still uncertain, existing studies provide some valuable insights. For example, the association of PA with inflammation has been systematically explored in previous work ^10^, which discovered that individuals engaging in regular PA had lower levels of C-reactive protein (CRP) and other inflammatory markers. Another study also suggested that PA could confer systemic beneficial modulations associated with inflammation ^11^. Simultaneously, CKD patients were commonly exposed to a chronic inflammatory state ^12^, which contributed to the progression of CKD ^13^; ^14^. Treatment approaches for CKD shifted towards inflammatory interventions, such as supplementing with probiotics, and also showed promise in alleviating the progression of CKD ^15^. Hence, we can reasonably hypothesize that inflammation might mediate the association of PA with CKD. Understanding this pathway is important to elaborate the biological mechanisms involved and to improve more precise interventions for CKD. To our knowledge, the mediating effect of inflammation on such association was not previously explored.

In the present study, we performed a mediation study by utilizing large-scale prospective data available from the UK Biobank ^16^. We aimed to investigate three important questions: (i) how PA affected the risk of CKD? To provide more robust evidence, we first examined the association of PA with incident CKD through PA data collected by the International Physical Activity Questionnaire (IPAQ) and accelerometers in two different sub-cohorts (questionnaire-measured cohort and accelerometer-measured cohort); (ii) how PA influenced the inflammatory status of the body? To achieve this, we assessed the association of PA with inflammation measured by four inflammation metrics including CRP, white blood cell (WBC), a low-grade inflammation (INFLA) score, and monocyte to high-density lipoprotein cholesterol ratio (MHR); (iii) Did there exist a mediating effect of inflammation standing on the path from PA to CKD? To this goal, we examined the mediating roles of inflammation in the association of PA with CKD.

## Methods and Materials

### Data sources and study populations

The UK Biobank is a large-scale prospective cohort, which recruited more than 500,000 participants aged 40-69 years between 2006 and 2010 in 22 assessment centers across the UK ^16^. The baseline information on social demography, function and physical measurements, and biochemical indexes were collected at enrollment.

To examine the association of PA with CKD and the mediation effects of inflammation on this association robustly, we generated two different sub-cohorts (Figure 1), including the questionnaire-measured cohort with 314,694 participants after removing participants with a diagnosis of CKD before recruitment (*N*=1,043) as well as those with missing PA and mediators (*N*=186,664), and the device-measured cohort with 79,095 participants (Figure S1). We conducted the primary analyses within the questionnaire-measured cohort and performed necessary supplementary analyses in the device-measured cohort to strengthen our conclusions obtained from the former.

**Figure 1.**
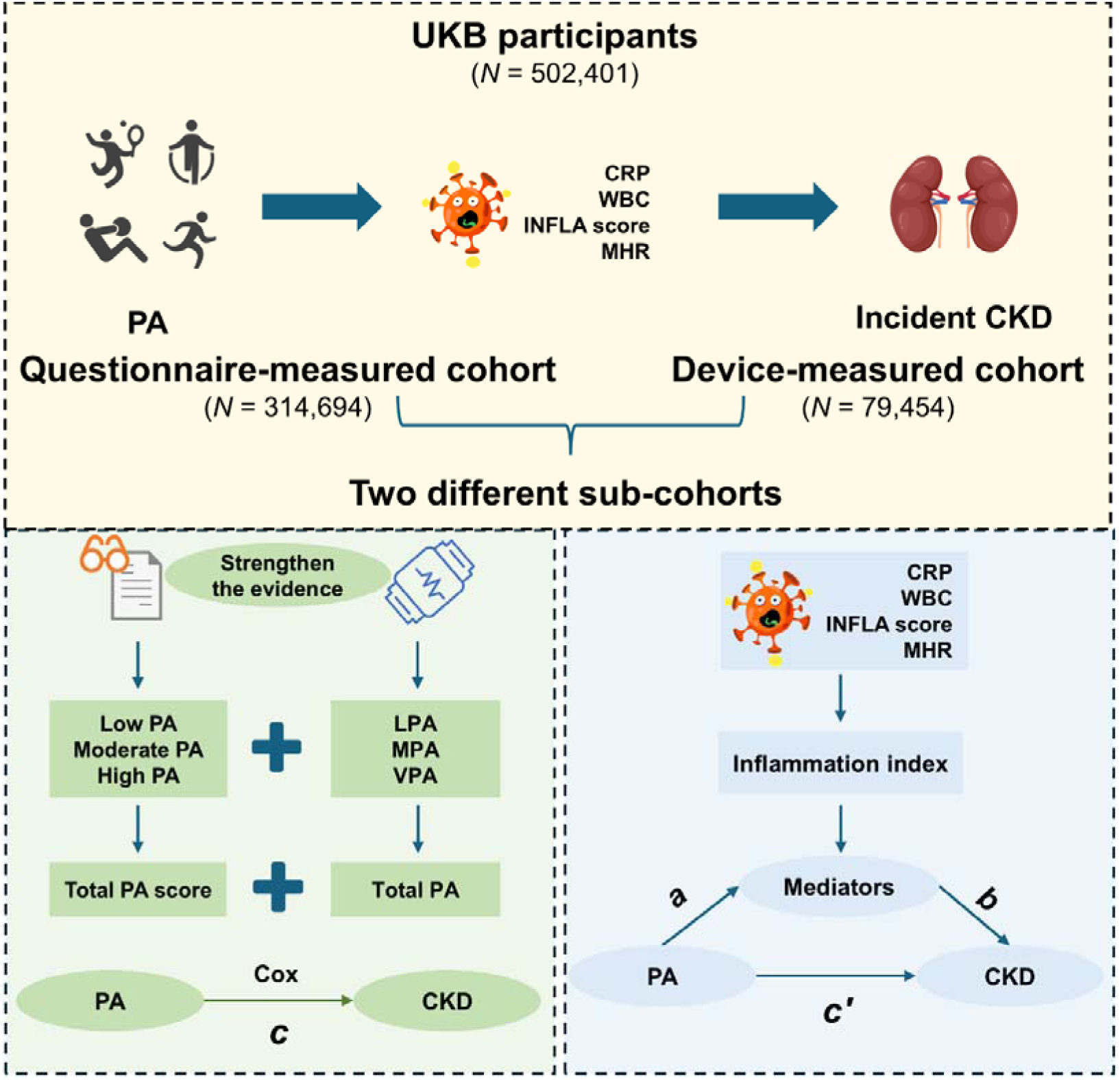
Flowchart of participant enrollment and data analysis in the present study. CKD: chronic kidney disease; PA: physical activity; CRP: C-reactive protein; WBC: white blood cell; INFLA score: a low-grade inflammation score; MHR: monocyte to high-density lipoprotein cholesterol ratio; *c* is the effect of PA on CKD; *c*_′_ is the effect of PA on CKD after controlling for the impact of inflammation; *a* is the effect of PA on inflammation; *b* is the effect of inflammation on CKD.

### Measurement of PA

#### Questionnaire-measured PA

The questionnaire-measured PA was assessed using the modified version of IPAQ, which captured the frequency and duration of walking, moderate, and vigorous-intensity activity ^17^. Weekly PA was represented via a composite score (MET-minutes/week), which was calculated as the sum of the minutes of walking, moderate-intensity, and vigorous-intensity activity every week, weighted by the metabolic equivalents (METs) for each type of activity according to the compilation given by ^18^. Note that, one MET was considered the metabolic rate generated during quiet sitting. Finally, following previous studies ^5^; ^19^, we categorized participants into three groups in terms of their weekly PA: low (<600 MET-min/week), moderate (600-3000 MET-min/week), and high (>3000 MET-min/week).

#### Device-measured PA

The device-measured PA was obtained by a wrist-mounted accelerometer ^20^. Participants were asked to wear the accelerometer for one week during their daily activities. Based on the captured triaxial acceleration data united in milligravities (mg), we calculated the time spent on 30-125 mg, 125-400 mg, and >400 mg, corresponding to light PA (LPA), moderate PA (MPA), and vigorous PA (VPA) ^21^, respectively. Then, referring to existing studies ^22^; ^23^, we transformed the time into energy expenditure by multiplying 3.3, 4.0, or 8.0, representing the average METs for LPA, MPA, and VPA, respectively. The total PA (TPA) was defined as the total energy expenditure of the above three PA types.

The data fields for calculating questionnaire-measured PA and device-measured PA are outlined in Table S1. In our data analyses, we standardized the continuous PA into *z*-score by dividing by the standard deviation (SD) after subtracting the mean.

### Diagnosis of CKD and determination of follow-up time

The primary outcome was incident CKD event, defined as the first diagnosis of CKD occurring during follow-up among participants free of CKD at baseline. Diagnoses obtained from hospital records and death registers (including England, Scotland, and Wales) ^16^, and recorded by the ICD-9 and ICD-10 coding systems (Table S2). The follow-up time for each participant was calculated from the recruitment date to the date of reporting a diagnostic CKD event, death, failure of follow-up, or follow-up deadline (19 July 2022), whichever occurred first.

### Covariate selection and missing value treatment

A wide range of potential covariates were considered, including age, sex (female or male), ethnicity (white or no white), drinking status (never, previous, or current drinker), smoking status (never, previous, or current smoker), Townsend deprivation index (TDI), education level (college or without college degree), income (<£31,000 or ≥£31,000) ^24^, and healthy diet score (ranging from 0 to 5) ^25^. Multiple imputation was performed by the chained equation multivariate estimation method to impute missing values for each covariate by R package “MICE” ^26^.

### Assessment of inflammation index

Two traditional inflammatory metrics, CRP and WBC, were selected to reflect the body’s inflammation level. ^27–29^. In addition to these individual metrics, the INFLA score and MHR were further calculated to assess the systemic inflammation. Specifically, the INFLA score was derived by combining the levels of four blood biomarkers (CRP, WBC, platelets, and neutrophil-to-lymphocyte ratio). Each biomarker was categorized into deciles based on its concentration: biomarkers in the top deciles (7 to 10) scored from 1 to 4, those in the bottom decile (1 to 4) ranging from -4 to -1, and those in the middle deciles (5 to 6) received a score of 0. Subsequently, the scores for the four biomarkers were aggregated to calculate the INFLA score, which ranged from -16 to 16, with higher scores indicating higher levels of inflammation ^30^. The MHR was defined as the monocyte count ratio to HDL-cholesterol ^29^. All inflammation-related metrics were standardized before analyses. Fields related to these metrics are displayed in Table S3.

### Statistical methods and data analyses

#### Baseline characteristics

We first summarized baseline characteristics in the two different sub-cohorts according to whether participants developed CKD in the follow-up. Baseline characteristics for continuous and categorical variables were expressed as mean ± SD or frequency (percentage), respectively. The between-group difference was compared using Student’s *t* or χ^2^ tests as appropriate. Additionally, we compared the baseline characteristics of participants with the development of CKD, stratified by physical activity levels: categorized as low, moderate, or high activity levels in the questionnaire-based cohort; and grouped by TPA in the device-measured cohort.

#### Construction of Cox models

In the questionnaire-measured cohort, three Cox models were established to assess the association of different levels of PA with incident CKD. Model 1 was a crude model without any covariates; Model 2 was adjusted for age, sex, and ethnicity; Model 3 was a fully adjusted model and further adjusted for smoking status, drinking status, TDI, education level, income, and healthy diet score. To strengthen the evidence for the association between PA and CKD, quartile-based stratification analyses for LPA, MPA, VPA, and TPA were also performed with the above Cox models in the device-measured cohort. Restricted cubic spline regressions were used to assess the nonlinear association between continuous PA and incident CKD in both cohorts. Hazard ratio (HR) and 95% confidence interval were reported.

#### Mediation analyses from PA to inflammation then to CKD

To determine the mediation effects of inflammation on the association of PA with CKD (Figure 1), we conducted a counterfactual mediation analysis in both two sub-cohorts : (i) Cox models were employed to examine the effect of PA on CKD (i.e., *c*), as discussed in the above section; (ii) linear regressions were applied to assess the effect of PA on inflammation (i.e., *a*); (iii) mediation models were constructed to evaluate both the effect of PA on CKD after controlling for the effect of inflammation as well as the influence of inflammation on CKD (i.e., *c*′ and *b*). Additionally, the proportion of the effect mediated and its corresponding 95% confidence intervals (CIs) were estimated through 5,000 bootstrap resamples. All analyses were adapted for the covariates incorporated in Model 3. ^31^.

#### Subgroup and sensitivity analyses

Due to the limited sample sizes of the device-measured cohort, subgroup and sensitivity analyses were only implemented within the questionnaire-measured cohort. To examine the consistency of the protective effect of PA across different covariate levels, we carried out several subgroup analyses stratified by age (<60 or ≥60 years), sex (female or male), education level (college or without college degree), income (<£31,000 or ≥£31,000), smoking status (never or ever), drink status (never or ever), and healthy diet score (0∼2 or 3∼5).

Considering the aging-related inflammation and the differences in inflammation status between females and males ^32^; ^33^, the mediation effects of inflammation were further validated across subgroups stratified by age and sex. In addition, we performed several sensitivity analyses: (i) to investigate the influence of missing values in covariates on the results ^34^, we repeated the analyses with participants having complete data; (ii) to minimize the reverse causality effect, we excluded participants who developed CKD during the first two years of follow-up; (iii) to eliminate the potential impact of certain diseases, we excluded participants with hypertension and diabetes at baseline; (iv) to control established risk factors by further adjusting for clinical histories of hypertension, diabetes, and obesity (body mass index ≥ 30 kg/m²).

#### Statistical software

All analyses were conducted with the R (version 4.3.0) software. The R MICE package was employed for missing value imputation ^26^, and the R CMAverse package was applied for the mediation analysis ^31^. Two-tailed *P* values<0.05 were considered statistically significant.

## Results

### Baseline characteristics for participants

The baseline characteristics of participants in the questionnaire-measured cohort are shown in Table 1. The average age was 56.8±8.1 years, 51.2% were female and 95.1% were of white ancestry. During a median follow-up time of 13.4 years, 10,676 CKD cases were identified, and were more likely to be older, male, have lower income, lower healthy diet score, lower TDI, active smokers, and previous drinkers. We also observed that CKD cases tended to have higher inflammation metrics. In the device-measured cohort, the average age of participants was 56.7±7.8 years, 56.0% were female and 96.7% were of white ancestry (Table S4). During a median follow-up time of 7.7 years, 1,594 CKD cases were identified. Similar baseline characteristics existed for participants in this cohort. Additionally, among participants diagnosed with CKD during follow-up, higher self-reported PA in the questionnaire cohort was associated with being older, male, non-smoking, higher educated, and having a healthier diet and lower inflammation (Table S5). However, some of the distinctions were not significant in the device-measured cohort when stratified by quartiles of TPA (Table S6).

**Table 1.**
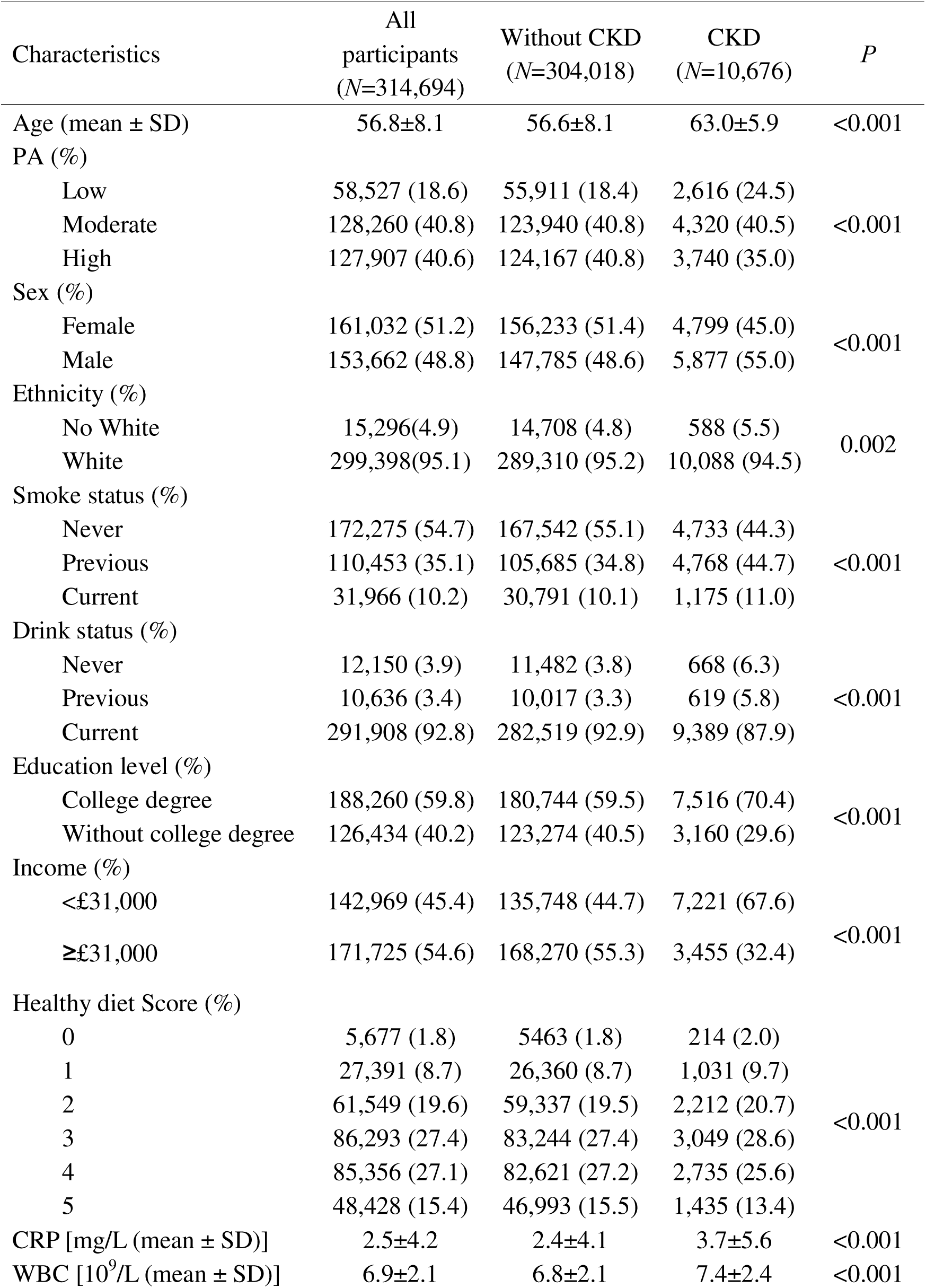
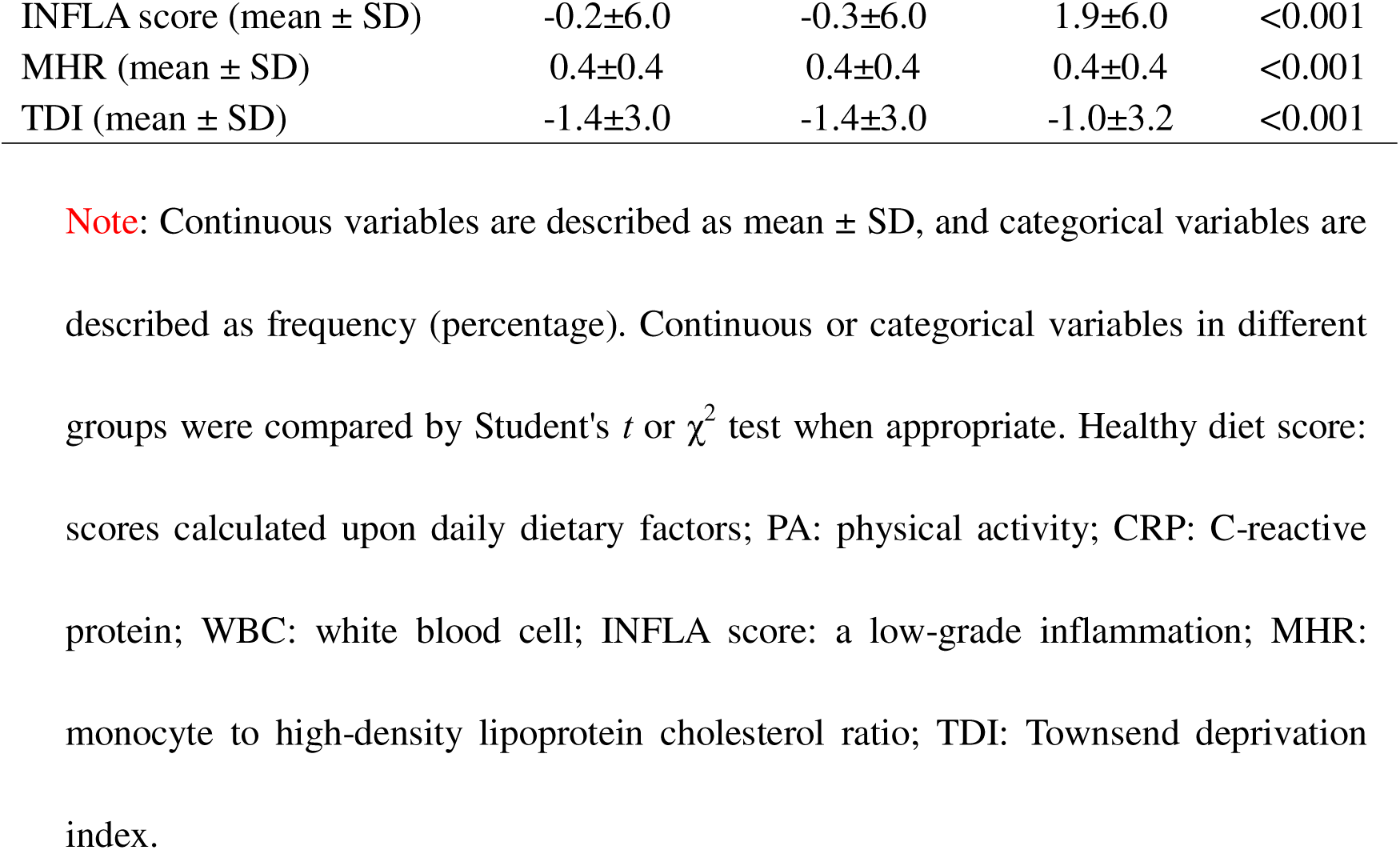
Baseline characteristics of all included participants of the questionnaire-measured cohort in the UK Biobank study.

### Association of PA with incident CKD

#### Association of PA with incident CKD in the questionnaire-measured cohort

In the questionnaire-measured cohort, a negatively correlated dose-response relation between total weekly PA and the risk of CKD was observed (Figure S2A). Notably, the protective effect of PA on CKD followed a threshold pattern, with the protective impact becoming apparent only when the total weekly PA dose reached approximately 1,700 MET minutes. Further, there existed significant differences in the cumulative risk of CKD among various PA intensity groups (*P*_Log-rank_<0.0001) in terms of the Kaplan-Meier curves (Figure S3).

After adjusting for confounding factors (Model 3), we found that, compared to low PA intensity, moderate and high PA intensity levels led to an approximately 28.0% (95% CI 24.4∼31.5%) and 37.6% (34.4∼40.7%) reduced risk of CKD, respectively; and higher PA level often tended to generate lower risk of incident CKD (*P*_trend_=6.25×10^-69^) (Figure 2). Similar association patterns regarding the protective impact of PA against CKD were also detected in model 1 and model 2.

**Figure 2.**
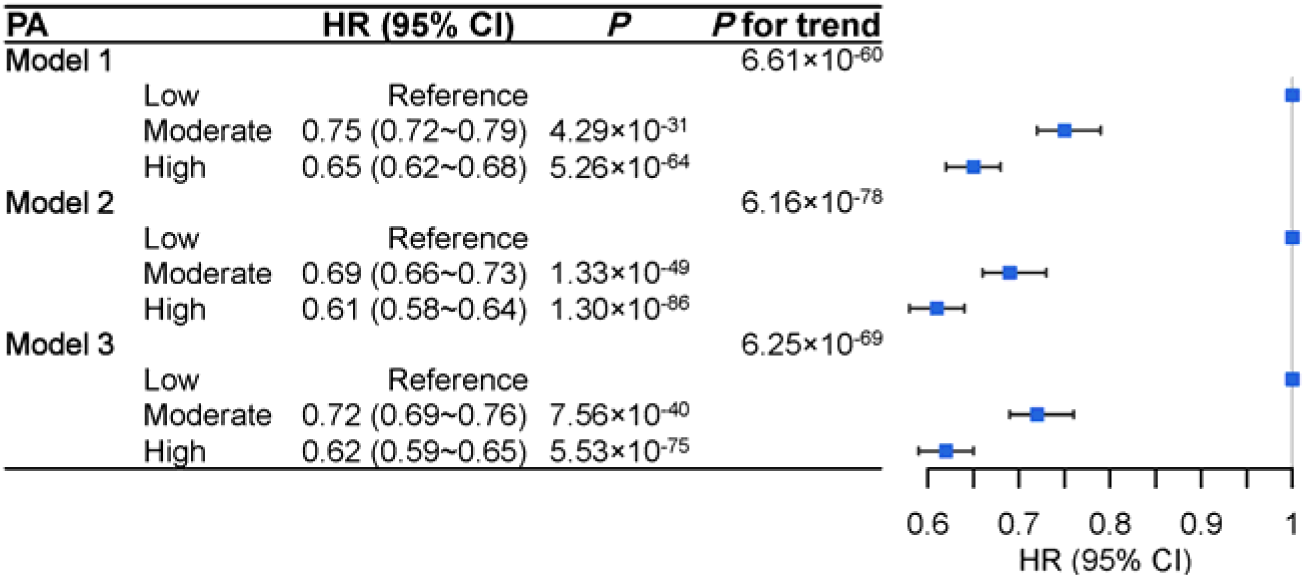
Association of PA with the risk of incident CKD in the questionnaire-measured cohort. PA: physical activity; CKD: chronic kidney disease; HR: hazard ratio; CI: confidence interval; Model 1: crude; Model 2: adjusted for age, sex, and ethnicity; Model 3: adjusted for age, sex, ethnicity, smoking status, drinking status, TDI, education, income, and healthy diet score.

#### Association of PA with incident CKD in the device-measured cohort

The protective effect of PA on CKD still existed in the device-measured cohort. The restricted cubic splines also showed a non-linear association between PA and the risk of CKD (Figure S2B-E), again showing a negative dose-response relation. The pattern of associations remained similar, indicating that a higher PA level was related to a lower risk of occurring CKD. Here, the minimal dose related to a decreased risk of CKD was around 1900 min/week, 420 min/week, 19 min/week, and 5500 MET/min/week for LPA, MPA, VPA, and TPA, respectively.

In terms of model 3, for per IQR increase in LPA, MPA, VPA, and TPA, the risk of developing CKD was reduced by approximately 16.6% (11.4∼21.5%), 39.0% (34.0∼43.6%), 6.6% (4.1∼9.1%), and 22.6% (17.8∼27.0%), respectively. Compared to Q1 (the lowest PA), the risk of occurring CKD for Q4 (the highest PA) decreased by 29.0% (19.3∼27.5%), 60.8% (54.3∼66.4%), 53.7% (45.9∼60.5%), and 37.2% (28.6∼45.7%) (Table 2), respectively. Similar results were also found in model 1 and model 2.

**Table 2.**
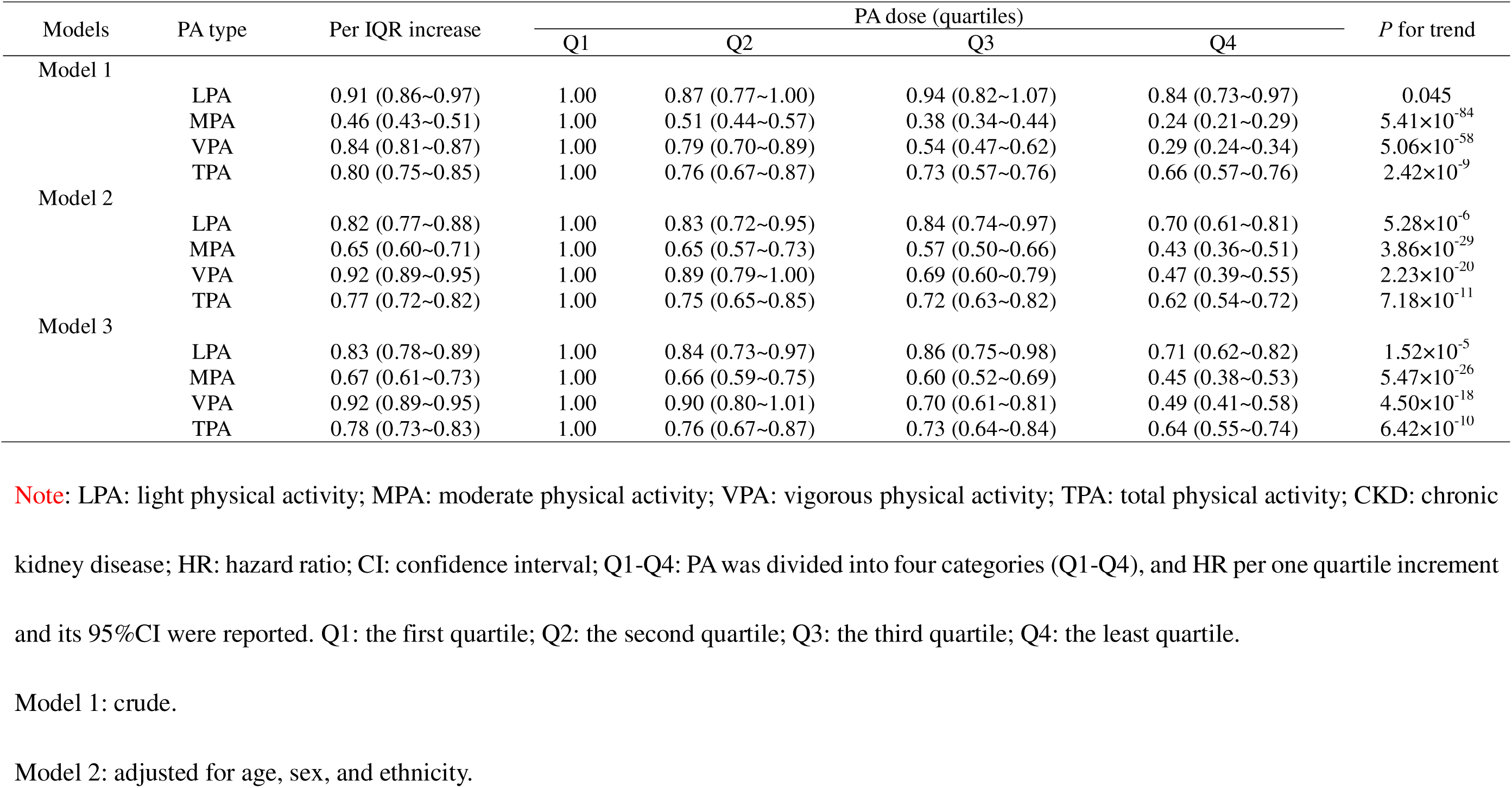
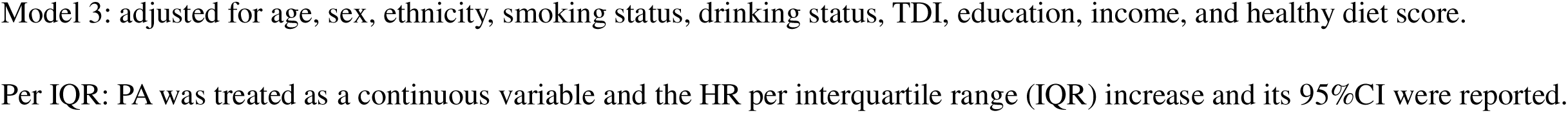
Association of LPA, MPA, VPA, and TPA with CKD in the device-measured cohort.

#### Subgroup and sensitivity analyses for the association of PA with CKD

In various subgroup analyses, PA remained robustly associated with a lower risk of incident CKD (Table S7). In particular, the protective effect of PA on CKD seemed to be slightly more pronounced among the former smokers and former drinkers as well as participants with higher education level, lower income, and more healthy diet. In various sensitivity analyses, consistent protection influences of PA against CKD were discovered when we repeated the analysis among participants with complete data (Table S8), or after removing participants with less than two years of follow-up (Table S9), or after excluding participants who had hypertension and diabetes at baseline (Table S10), or after further taking the clinical histories of hypertension, diabetes, and obesity into account (Table S11).

### Mediating path from PA to inflammation then to CKD

#### Estimated effects of PA on inflammation

In the questionnaire-measured cohort, we observed that PA displayed a significantly negative correlation with various inflammation metrics. Specifically, the estimated effect was -0.051 (*se*=0.002), -0.038 (*se*=0.002), -0.056 (*se*=0.002), and -0.036 (*se*=0.002) for CRP, WBC, INFLA score and MHR, respectively. Notably, PA exhibited the strongest association with the INFLA score, followed by CRP among all the inflammation metrics (Table 3).

**Table 3.**
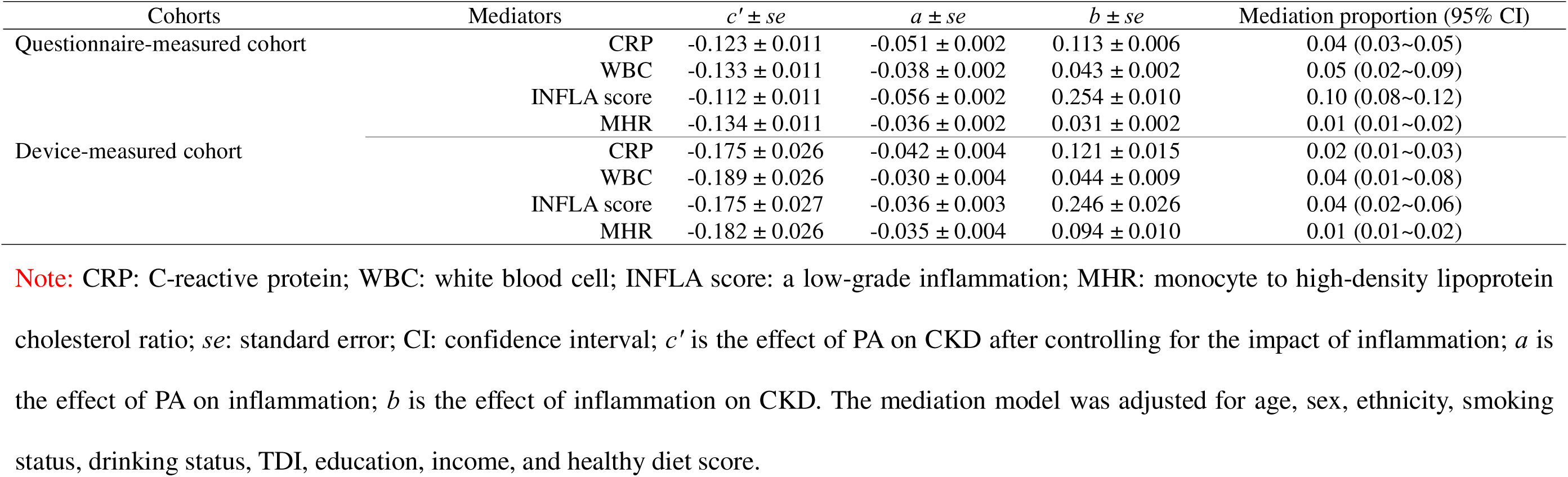
Mediation analyses of inflammation in two different sub-cohorts in the UK Biobank study.

In the device-measured cohort, the negative effects of PA on inflammation remained evident, but were slightly decreased compared to those obtained in the questionnaire-measured cohort. The estimated effects were -0.042 (*se*=0.004), -0.030 (*se*=0.004), -0.036 (*se*=0.003), and -0.035 (*se*=0.004) for CRP, WBC, INFLA score, and MHR, respectively (Table 3).

#### Association of inflammation with incident CKD

In the questionnaire-measured cohort, the risk of CKD events increased by 11.2% (10.6∼13.2%), 4.4% (3.9∼4.9%), 29.0% (26.4∼31.5%), and 3.1% (2.7∼3.5%) for per SD increment in CRP, WBC, INFLA score, and MHR, respectively (Table 3). Similar effects of inflammation on occurring CKD were observed in the device-measured cohort, with the risk increased by 11.8% (9.7∼16.1%), 4.5% (2.7∼6.3%), 27.9% (21.5∼34.6%), and 9.9% (7.6∼12.3%) or per SD increment in CRP, WBC, INFLA score, and MHR (Table 3), respectively.

#### Mediating roles of inflammation in the association of PA and CKD

In the questionnaire-measured cohort, we discovered that inflammation substantially mediated the effect of PA on CKD (Figure 3A-D and Table 3). Particularly, the INFLA score emerged as the primary factor mediating the association of PA with CKD among the inflammation metrics, respectively. The mediation proportions were 4.1% (3.0∼7.7%) 1.4% (1.1∼1.7%), 9.8% (7.7∼11.9%), and 1.4% (1.1∼1.7%) for CRP, WBC, INFLA score and MHR, respectively.

**Figure 3.**
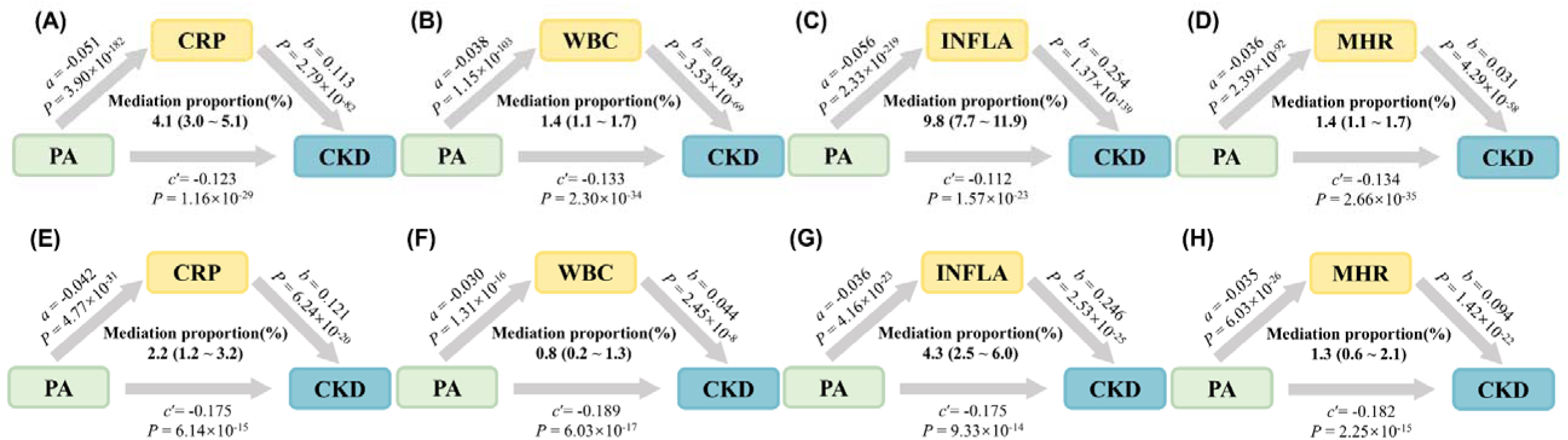
(A-D) Mediating role of inflammation in the association of PA with CKD in the questionnaire-measured cohort. (E-H) Mediating role of inflammation in the association of PA with CKD in the device-measured cohort. PA: physical activity; CKD: chronic kidney disease; CRP: C-reactive protein; WBC: white blood cell; INFLA score: a low-grade inflammation; MHR: monocyte to high-density lipoprotein cholesterol ratio; *c*_′_ is the effect of PA on CKD after controlling for the impact of inflammation; *a* is the effect of PA on inflammation; *b* is the effect of inflammation on CKD. The mediation model was adjusted for age, sex, ethnicity, smoking status, drinking status, TDI, education, income, and healthy diet score.

When comes to the device-measured cohort, results suggested that the magnitude of the effects was somewhat attenuated, with the mediation proportions being 2.2% (1.2∼3.2%), 0.8% (0.2∼1.3%), 4.3% (2.5∼6.0%), and 1.3% (0.6∼2.1%) for CRP, WBC,

INFLA score and MHR, respectively (Figure 3E-H and Table 3). Further, we needed to highlight that we did not detect any significant interaction effects between PA and inflammation metrics on the risk of encountering CKD.

#### Subgroup and sensitivity analysis for the mediating roles of inflammation

In subgroup analyses, the mediating role of inflammation remained significant across sex or age groups (Tables S12-S13). In the sensitivity analyses, inflammation played a similar mediating role when we repeated the analysis among participants with complete data (Table S14). After excluding CKD cases occurring with less than two years of follow-up, the mediation effects of inflammation remained significant (Table S15). After removing participants who had hypertension and diabetes at baseline, similar mediation effects of inflammation were observed (Table S16). The mediation effects of inflammation remained unchanged appreciably after further explaining the clinical history of hypertension, diabetes, and obesity (Table S17).

## Discussion

### Summary of our study

In this large prospective cohort study, we conducted a comprehensive analysis to investigate the association between PA and incident CKD. In the questionnaire-measured cohort, we found that higher PA intensity was associated with a reduced risk of developing CKD. Using objective data from the device-measured cohort, we obtained consistent results and validated the dose-response relationships between different PA intensities (LPA, MPA, VPA) and CKD incidence. Furthermore, we observed that PA was significantly inversely related to inflammation, which partially mediated the association between PA and CKD. Notably, the INFLA score emerged as the strongest mediator among the inflammation metrics.

### Comparison to previous studies

#### Comparison with previous studies on the association of PA with CKD

The association between PA and CKD discovered in this work was broadly consistent with and complementary to previous studies. For example, a prior meta-analysis showed that PA could reduce the incidence of CKD to some extent ^8^; several prospective studies also supported this observation ^5^; ^7^. However, not all existing studies confirmed the protective effect of PA on CKD. A study involving 3,075 well-functioning older adults discovered that PA was likely related to a faster decline in kidney function and a higher risk of incident CKD ^35^, similar result was found in an occupational cohort in Japan ^36^. Limited sample sizes and special population groups they chose might probably account for those inconsonant observations. Meanwhile, we discovered the minimal dose for the protective effect of PA on CKD, which also existed for other diseases according to a previous study ^37^.

#### Comparison with previous studies on inflammation metrics

Substantial evidence supported that regular PA could be an effective anti-inflammatory measure ^11^, which was consistent with our results. A recent study performed in two independent prospective cohorts (*n*=142, a median follow-up of 6.5 years; *n*=103, a median follow-up of 8.0 years) examined the relations between multiple inflammation-related biomarkers (e.g., IL-8) and CKD, and revealed that nine were significantly associated with the incidence of CKD and renal damage ^38^. However, to our knowledge, we are the first to comprehensively examine the mediating role of inflammation in the association between PA and CKD, thereby bridging existing research gaps.

Moreover, prior studies have generally focused on the single inflammation indicator, which failed to fully represent the systemic inflammatory state ^39^. In contrast, our study incorporated two traditional inflammation markers as well as two composite inflammation indices. As hypothesized, our findings confirmed that inflammation significantly mediated the association of PA and CKD events, with the INFLA score exhibiting the most pronounced mediating effect.

### Possible biological mechanisms

The mediating role of inflammation in the association of PA with CKD might be attributed to the following mechanisms. The damage to the glomerular capillaries and elevated oxidative stress levels induced by inflammation might explain the mediating role of inflammation, which ultimately led to a decline in the glomerular filtration rate ^40^. Failure to promptly repair this damage might culminate in renal tissue fibrosis, resulting in irreversible harm ^41^. All these were associated with an increased risk of incident CKD ^4^. PA increased the activity of antioxidant enzymes and reduced oxygen free radicals, thus reducing these damages ^33^.

### Public health implications

Our study has some public health implications. First, our findings indicated PA was associated with a reduced risk of developing CKD, which provided a scientific basis for developing prevention strategies. By encouraging people to be physically active regularly, the risk of developing CKD could be reduced, which will contribute to the reduction of the burden on patients and society ^1^.

Second, we identified the mediating roles of inflammation in the association between PA with CKD, which indicated that interventions targeting inflammation, such as inhibiting inflammatory pathways ^15^, might enhance the protective effects of PA against CKD.

Third, our study confirmed that the INFLA score exhibited the most pronounced mediating role in the association between PA and CKD, suggesting that the future development of such comprehensive inflammatory indicators could be more beneficial in elucidating health-related outcomes.

### Advantages of our study

Our study has several advantages. First, the UK Biobank contained detailed participant information, allowing for a range of confounding factor adjustments in statistical analyses. The large sample sizes of the UK Biobank also offered the basis for subgroup analysis and sensitivity analysis, which significantly enhanced the robustness of our research findings. Second, our prospective study had a relatively long follow-up time, which reduced the effect of reverse causality. Finally, we further utilized accelerometer-measured PA to strengthen the evidence on the association of PA with CKD. This approach accurately measured the intensity and duration of PA throughout the wearing period and circumvented recall bias and social desirability bias induced by self-report ^42^.

### Limitations of our study

Our study also has several limitations. First, PA obtained from questionnaires may lead to recall bias and social desirability bias ^42^. Second, although PA obtained from accelerometers might be more objective, certain types of activities, such as resistance exercise or cycling, may not be reflected by the accelerometers ^9^. Third, the baseline PA we used did not take the changes in PA intensity over the follow-up time into account. Data from a follow-up study showed that PA intensities stabilized over time, with a slight decline in time spent in high PA intensity ^43^. Therefore, our findings might slightly underestimate the association between PA and incident CKD. Finally, because of the very limited sample size of the follow-up data, we had to analyze PA and the mediators (e.g., CPR) measured at baseline, which may lead to the concern of reverse causality between PA and these mediators. However, as shown in previous work that the correlations of these mediators measured at baseline and the three follow-ups were considerably high ^22^, the similar relations between PA and these mediators should still maintain if we analyzed sufficient follow-up samples. In addition, the reverse influence of the mediators on PA seemed not be reasonable from a biological perspective. These suggested the reverse causality bias was minimal on our mediation analysis in the device-measured cohort.

### Conclusions

This study shows suggestive evidence for the association of active PA with reduced risk of CKD events and further reveals a potential mediating role of inflammation in such an association, providing a novel perspective for the early prevention of CKD.

## Data Availability

This study used the UK Biobank resource with the application ID 88159. Researchers can access to the UK Biobank dataset by applying to the UK Biobank official website (https://www.ukbiobank.ac.uk). All data generated or analyzed during this study are included in this published article and its supplementary information files.

## Additional File

Supplementary Files.

## Abbreviations

CKD: chronic kidney disease
PA: physical activity
CRP: C-reactive protein
WBC: white blood cell
INFLA: low-grade inflammation
MHR: monocyte-to-high density lipoprotein-cholesterol ratio
IPAQ: International Physical Activity Questionnaire
MET: metabolic equivalent
LPA: light physical activity
MPA: moderate physical activity
VPA: vigorous physical activity
TPA: total physical activity

## Acknowledgements

The UK Biobank approval was given for this project under the application number of 88159 and can be downloaded from https://www.UK Biobankiobank.ac.uk/ or https://biota.osc.ox.ac.uk/. The data analyses in the present study were carried out with the high-performance computing cluster that was supported by the special central finance project of local universities for Xuzhou Medical University. We thank the Editor, Associate Editor, and anonymous reviewers for their important and constructive comments, which substantially improved our manuscript.

## Author Contributions

PZ conceived the idea for the study. PZ obtained the data. PZ and XZ cleared up the datasets, performed the data analyses, interpreted the results of the data analyses and wrote the manuscript.

## Availability of data and materials

This study used the UK Biobank resource with the application ID 88159. Researchers can access to the UK Biobank dataset by applying to the UK Biobank official website (https://www.UK Biobankiobank.ac.uk/). All data generated or analyzed during this study are included in this published article and its supplementary information files.

## Ethics approval and consent to participate

The UK Biobank had approval from the North West Multi-Centre Research Ethics Committee (MREC) as a Research Tissue Bank (RTB) approval. All participants provided written informed consent before enrolment in the study, which was conducted in accordance with the Declaration of Helsinki. This approval means that other researchers do not require separate ethical clearance and can operate under the RTB approval.

## Consent for publication

Not applicable.

