## Supplementary_File for "Mediation effects of inflammation on the association of physical activity and chronic kidney disease: evidence from questionnaire and device-measured assessments"

Table S1. Fields related to the measurement of three specific types of physical activity in the questionnaire-measured cohort and device-measured cohort of the UK Biobank study.

|  | Physical activity types | Fields |
| --- | --- | --- |
| Questionnaire-measured cohort | | |
|  | Walking | 864, 874 |
|  | Moderate activity | 884, 894 |
|  | Vigorous activity | 904, 914 |
| Device-measured cohort | | |
|  | LPA | 90113, 90128 |
|  | MPA | 90128, 90139 |
|  | VPA | 90139 |

Note: LPA: light physical activity; MPA: moderate physical activity; VPA: vigorous physical activity.

Table S2. Definitions and sources of information for CKD and other diseases in the UK Biobank study.

| Disease | ICD-9 | ICD-10 |
| --- | --- | --- |
| CKD | 585, 5859 | I12.0, I13.1, I13.2, N18, N18.0, N18.2, N18.3, N18.4, N18.5, N18.8, N18.9 |
| Hypertension | 401, 4010, 4011, 4019, 402, 4020, 4021, 4029, 403, 4030, 4031, 4039, 404, 4040, 4041, 4049, 405, 4050, 4051, 4059 | I10, I11, I11.0, I11.9, I12, I12.0, I12.9, I13, I13.0, I13.1, I13.2, I13.9, I15, I15.0, I15.1, I15.2, I15.8, I15.9 |
| Diabetes | 249, 250 | E10, E11, E12, E13, E14, I64 |

Note: ICD: International Classification of Disease; CKD: chronic kidney disease.

Table S3. Fields related to the inflammation metrics in the UK Biobank study.

| Metric | Relevant variables used | Field |
| --- | --- | --- |
| CPR (mg/L) | CRP | 30710 |
| WBC (10^9^/L) | WBC | 30000 |
| INFLA score | CRP | 30710 |
|  | WBC | 30000 |
|  | Platelets | 30080 |
|  | Neutrophil | 30140 |
|  | Lymphocyte | 30120 |
| MHR | HDL-C | 30760 |
|  | Monocyte | 30130 |

Note: CRP: C-reactive protein; WBC: white blood cell; INFLA score: a low-grade inflammation score; MHR: monocyte to high-density lipoprotein cholesterol ratio.

Table S4. Baseline characteristics of all included participants of the device-measured cohort in the UK Biobank study.

| Characteristics | All participants  (*N*=79,095) | Without CKD  (*N*=77 501) | Incident CKD  (*N*=1 594) | *P* |
| --- | --- | --- | --- | --- |
| Age (mean ± SD) | 56.7±7.8 | 56.6±7.8 | 62.6±5.8 | <0.001 |
| Sex (%) | | | | |
| Female | 44,150 (55.8) | 43,396 (56.0) | 754 (47.3) | <0.001 |
| Male | 34,945 (44.2) | 34,105 (44.0) | 840 (52.7) |  |
| Ethnicity (%) | | | | |
| No White | 2,609 (3.30) | 2,551 (3.29) | 58 (3.64) | 0.486 |
| White | 76,486 (96.7) | 74,950 (96.7) | 1,536 (96.4) |  |
| Smoke status (%) | | | | |
| Never | 45,128 (57.1) | 44,335 (57.2) | 793 (49.7) | <0.001 |
| Previous | 28,476 (36.0) | 27,793 (35.9) | 683 (42.8) |  |
| Current | 5,491 (6.94) | 5,373 (6.93) | 118 (7.40) |  |
| Drink status (%) | | | | |
| Never | 2,223 (2.81) | 2,151 (2.78) | 72 (4.52) | <0.001 |
| Previous | 2,132 (2.70) | 2,057 (2.65) | 75 (4.71) |  |
| Current | 74,740 (94.5) | 73,293 (94.6) | 1,447 (90.8) |  |
| Education level (%) | | | | |
| College | 41,959 (53.0) | 40,917 (52.8) | 1,042 (65.4) | <0.001 |
| Without college degree | 37,136 (47.0) | 36,584 (47.2) | 552 (34.6) |  |
| Income (%) | | | | |
| <£31 000 | 29,869 (37.8) | 29,038 (37.5) | 831 (52.1) | <0.001 |
| ≥£31 000 | 49,226 (62.2) | 48,463 (62.5) | 763 (47.9) |  |
| Healthy diet score (%) | | | | |
| 0 | 1,092 (1.38) | 1,076 (1.39) | 16 (1.00) | 0.022 |
| 1 | 5,919 (7.48) | 5,778 (7.46) | 141 (8.85) |  |
| 2 | 14,142 (17.9) | 13,822 (17.8) | 320 (20.1) |  |
| 3 | 21,878 (27.7) | 21,443 (27.7) | 435 (27.3) |  |
| 4 | 22,754 (28.8) | 22,319 (28.8) | 435 (27.3) |  |
| 5 | 13,310 (16.8) | 13,063 (16.9) | 247 (15.5) |  |
| LPA [min/week (mean ± SD)] | 5,206±1,312 | 5,208±1,312 | 5,117±1,330 | 0.007 |
| MPA [min/week (mean ± SD)] | 385±318 | 388±319 | 253±251 | <0.001 |
| VPA [min/week (mean ± SD)] | 7.50±31.5 | 7.61±31.7 | 2.43±9.00 | <0.001 |
| TPA [MET/min/week (mean ± SD)] | 5,599±1,321 | 5,604±1,320 | 5,373±1,365 | <0.001 |
| CRP [mg/L (mean ± SD)] | 2.24±3.87 | 2.21±3.83 | 3.47±5.45 | <0.001 |
| WBC [10^9^/L (mean ± SD)] | 6.68±1.93 | 6.68±1.93 | 7.13±1.86 | <0.001 |
| INFLA score (mean ± SD) | -0.70±5.94 | -0.73±5.93 | 1.08±5.88 | <0.001 |
| MHR (mean ± SD) | 0.34±0.19 | 0.34±0.19 | 0.40±0.26 | <0.001 |
| TDI (mean ± SD) | -1.7±2.8 | -1.74±2.81 | -1.74±2.88 | 0.999 |

Note: Continuous variables are described as mean ± SD, and categorical variables are described as frequency (percentage). Continuous or categorical variables in different groups were compared by Student′s *t* or χ^2^ test when appropriate. Healthy diet score: scores calculated upon daily dietary factors; LPA: light physical activity; MPA: moderate physical activity; VPA: vigorous physical activity; TPA: total physical activity; CRP: C-reactive protein; WBC: white blood cell; INFLA score: a low-grade inflammation score; MHR: monocyte to high-density lipoprotein cholesterol ratio; TDI: Townsend deprivation index.

Table S5. Baseline characteristics of low, moderate, and high physical activity patients in the questionnaire cohort.

| Characteristics | All patients  (*N*=10,676) | Low PA  (*N*=2,616) | Moderate PA  (*N*=4,320) | High PA  (*N*=3,740) | *P* |
| --- | --- | --- | --- | --- | --- |
| Age (mean ± SD) | 63.0 (5.94) | 62.6 (5.98) | 63.1 (5.87) | 63.1 (5.99) | <0.001 |
| Sex (%) | | | | | <0.001 |
| Female | 4,799 (45.0) | 1,170 (44.7) | 2,043 (47.3) | 1,586 (42.4) |  |
| Male | 5,877 (55.0) | 1,446 (55.3) | 2,277 (52.7) | 2,154 (57.6) |  |
| Ethnicity (%) | | | | | 0.124 |
| No White | 588 (5.51) | 153 (5.85) | 252 (5.83) | 183 (4.89) |  |
| White | 10,088 (94.5) | 2,463 (94.2) | 4,068 (94.2) | 3,557 (95.1) |  |
| Smoke status (%) | | | | | 0.007 |
| No | 4,733 (49.8) | 1,065 (47.0) | 1,989 (51.1) | 1,679 (50.2) |  |
| Yes | 4,768 (50.2) | 1,201 (53.0) | 1,902 (48.9) | 1,665 (49.8) |  |
| Drink status (%) | | | | | 0.458 |
| No | 668 (51.9) | 193 (52.0) | 240 (49.9) | 235 (54.0) |  |
| Yes | 619 (48.1) | 178 (48.0) | 241 (50.1) | 200 (46.0) |  |
| Education level (%) | | | | | <0.001 |
| College degree | 7,516 (70.4) | 1,816 (69.4) | 2,978 (68.9) | 2,722 (72.8) |  |
| Without college degree | 3,160 (29.6) | 800 (30.6) | 1,342 (31.1) | 1,018 (27.2) |  |
| Income (%) | | | | | 0.216 |
| <£31,000 | 7,221 (67.6) | 1,781 (68.1) | 2,881 (66.7) | 2,559 (68.4) |  |
| ≥£31,000 | 3,455 (32.4) | 835 (31.9) | 1,439 (33.3) | 1,181 (31.6) |  |
| Healthy diet Score (%) | | | | | <0.001 |
| 0 | 214 (2.00) | 74 (2.83) | 82 (1.90) | 58 (1.55) |  |
| 1 | 1,031 (9.66) | 339 (13.0) | 388 (8.98) | 304 (8.13) |  |
| 2 | 2,212 (20.7) | 609 (23.3) | 908 (21.0) | 695 (18.6) |  |
| 3 | 3,049 (28.6) | 777 (29.7) | 1,207 (27.9) | 1,065 (28.5) |  |
| 4 | 2,735 (25.6) | 570 (21.8) | 1,144 (26.5) | 1,021 (27.3) |  |
| 5 | 1,435 (13.4) | 247 (9.44) | 591 (13.7) | 597 (16.0) |  |
| CRP [mg/L (mean ± SD)] | 3.74±5.55 | 4.67±6.40 | 3.56±5.27 | 3.30±5.15 | <0.001 |
| WBC [10^9^/L (mean ± SD)] | 7.39±2.35 | 7.72±2.87 | 7.33±2.00 | 7.24±2.29 | <0.001 |
| INFLA score (mean ± SD) | 1.85±6.01 | 2.95±6.02 | 1.70±6.03 | 1.25±5.88 | <0.001 |
| MHR (mean ± SD) | 0.44±0.44 | 0.48±0.38 | 0.43±0.26 | 0.43±0.61 | <0.001 |
| TDI (mean ± SD) | -1.02±3.23 | -0.77±3.31 | -1.08±3.24 | -1.13±3.14 | <0.001 |

Note: Continuous variables are described as mean ± SD, and categorical variables are described as frequency (percentage). Continuous or categorical variables in different groups were compared by Student′s *t* or χ^2^ test when appropriate. Healthy diet score: scores calculated upon daily dietary factors; CRP: C-reactive protein; WBC: white blood cell; INFLA score: a low-grade inflammation score; MHR: monocyte to high-density lipoprotein cholesterol ratio; TDI: Townsend deprivation index.

Table S6. Baseline characteristics of participants who developed CKD stratified by quartiles of total physical activity in the device-measured cohort.

| Characteristics | All patients  (*N*=1,594) | Quartile 1  (*N*=399) | Quartile 2  (*N*=398) | Quartile 3  (*N*=398) | Quartile 4  (*N*=399) | *P* |
| --- | --- | --- | --- | --- | --- | --- |
| Age (mean ± SD) | 62.6 (5.76) | 62.5 (5.62) | 62.8 (6.18) | 62.7 (5.67) | 62.5 (5.54) | 0.876 |
| Sex (%) | | | | | | <0.001 |
| Female | 754 (47.3) | 138 (34.6) | 174 (43.7) | 202 (50.8) | 240 (60.2) |  |
| Male | 840 (52.7) | 261 (65.4) | 224 (56.3) | 196 (49.2) | 159 (39.8) |  |
| Ethnicity (%) | | | | | | 0.405 |
| No White | 58 (3.64) | 12 (3.01) | 13 (3.27) | 13 (3.27) | 20 (5.01) |  |
| White | 1,536 (96.4) | 387 (97.0) | 385 (96.7) | 385 (96.7) | 379 (95.0) |  |
| Smoke status (%) | | | | | | 0.708 |
| No | 793 (53.7) | 189 (53.7) | 209 (55.7) | 202 (54.0) | 193 (51.5) |  |
| Yes | 683 (46.3) | 163 (46.3) | 166 (44.3) | 172 (46.0) | 182 (48.5) |  |
| Drink status (%) | | | | | | 0.333 |
| No | 72 (49.0) | 22 (52.4) | 16 (51.6) | 12 (35.3) | 22 (55.0) |  |
| Yes | 75 (51.0) | 20 (47.6) | 15 (48.4) | 22 (64.7) | 18 (45.0) |  |
| Education level (%) | | | | | | 0.710 |
| College degree | 1,042 (65.4) | 255 (63.9) | 267 (67.1) | 255 (64.1) | 265 (66.4) |  |
| Without college degree | 552 (34.6) | 144 (36.1) | 131 (32.9) | 143 (35.9) | 134 (33.6) |  |
| Income (%) | | | | | | 0.576 |
| <£31,000 | 831 (52.1) | 219 (54.9) | 204 (51.3) | 208 (52.3) | 200 (50.1) |  |
| ≥£31,000 | 763 (47.9) | 180 (45.1) | 194 (48.7) | 190 (47.7) | 199 (49.9) |  |
| Healthy diet Score (%) | | | | | | <0.001 |
| 0 | 16 (1.00) | 7 (1.75) | 5 (1.26) | 3 (0.75) | 1 (0.25) |  |
| 1 | 141 (8.85) | 45 (11.3) | 30 (7.54) | 43 (10.8) | 23 (5.76) |  |
| 2 | 320 (20.1) | 92 (23.1) | 78 (19.6) | 72 (18.1) | 78 (19.5) |  |
| 3 | 435 (27.3) | 108 (27.1) | 127 (31.9) | 111 (27.9) | 89 (22.3) |  |
| 4 | 435 (27.3) | 103 (25.8) | 99 (24.9) | 106 (26.6) | 127 (31.8) |  |
| 5 | 247 (15.5) | 44 (11.0) | 59 (14.8) | 63 (15.8) | 81 (20.3) |  |
| CRP [mg/L (mean ± SD)] | 3.47 (5.45) | 4.31 (7.34) | 3.30 (5.15) | 3.29 (4.30) | 2.98 (4.37) | 0.004 |
| WBC [10^9^/L (mean ± SD)] | 7.13 (1.86) | 7.43 (2.10) | 7.05 (1.85) | 7.01 (1.76) | 7.01 (1.68) | 0.002 |
| INFLA score (mean ± SD) | 1.08 (5.88) | 1.72 (6.21) | 0.80 (5.70) | 0.99 (5.77) | 0.83 (5.79) | 0.092 |
| MHR (mean ± SD) | 0.40 (0.26) | 0.44 (0.22) | 0.42 (0.33) | 0.40 (0.27) | 0.36 (0.18) | <0.001 |
| TDI (mean ± SD) | -1.74 (2.88) | -1.10 (3.11) | -1.86 (2.88) | -1.88 (2.82) | -2.12 (2.60) | <0.001 |

Note: Continuous variables are described as mean ± SD, and categorical variables are described as frequency (percentage). Continuous or categorical variables in different groups were compared by Student′s *t* or χ^2^ test when appropriate. CKD: chronic kidney disease; Healthy diet score: scores calculated upon daily dietary factors; CRP: C-reactive protein; WBC: white blood cell; INFLA score: a low-grade inflammation score; MHR: monocyte to high-density lipoprotein cholesterol ratio; TDI: Townsend deprivation index.

Table S7. Subgroup analysis for the association of PA with incident CKD.

| Subgroup | N | PA level | | |
| --- | --- | --- | --- | --- |
|  |  | Low | Moderate | High |
| Sex |  |  |  |  |
| Female | 161,032 | 1.00 | 0.70 (0.66~0.76) | 0.60 (0.56~0.65) |
| Male | 153,662 | 1.00 | 0.73 (0.69~0.78) | 0.62 (0.58~0.67) |
| Ethnicity |  |  |  |  |
| Not White | 15,296 | 1.00 | 0.85 (0.70~1.05) | 0.67 (0.54~0.83) |
| White | 299,398 | 1.00 | 0.71 (0.68~0.75) | 0.61 (0.58~0.64) |
| Age |  |  |  |  |
| <60 | 182,257 | 1.00 | 0.71 (0.64~0.78) | 0.59 (0.53~0.65) |
| ≥60 | 132,47 | 1.00 | 0.72 (0.69~0.77) | 0.62 (0.59~0.66) |
| Smoking status |  |  |  |  |
| No | 172,275 | 1.00 | 0.76 (0.71~0.84) | 0.65 (0.60~0.70) |
| Yes | 142,419 | 1.00 | 0.69 (0.65~0.73) | 0.59 (0.55~0.63) |
| Drinking status |  |  |  |  |
| No | 12,150 | 1.00 | 0.66 (0.55~0.80) | 0.71 (0.59~0.86) |
| Yes | 302,544 | 1.00 | 0.72 (0.69~0.76) | 0.61 (0.58~0.64) |
| Education |  |  |  |  |
| Without College degree | 188,260 | 1.00 | 0.73 (0.69~0.78) | 0.63 (0.59~0.67) |
| College degree | 171,725 | 1.00 | 0.69 (0.63~0.76) | 0.58 (0.53~0.64) |
| Income |  |  |  |  |
| <£31000 | 142,969 | 1.00 | 0.69 (0.65~0.74) | 0.59 (0.55~0.62) |
| ≥£31000 | 171,725 | 1.00 | 0.78 (0.71~0.85) | 0.68 (0.62~0.74) |
| Healthy diet score | |  |  |  |
| 0-2 | 94,617 | 1.00 | 0.75 (0.69~0.81) | 0.67 (0.61~0.72) |
| 3-5 | 220,077 | 1.00 | 0.70 (0.66~0.75) | 0.59 (0.55~0.63) |

Note: Healthy diet score: scores calculated upon daily dietary factors; PA: physical activity; CKD: chronic kidney disease.

Table S8. Association of PA with CKD among participants with complete covariate data.

| PA | Model 1 | | Model 2 | | Model 3 | |
| --- | --- | --- | --- | --- | --- | --- |
|  | HR (95% CI) | *P* | HR (95% CI) | *P* | HR (95% CI) | *P* |
| Low | Reference | | Reference | | Reference | |
| Moderate | 0.80 (0.75~0.85) | 1.78×10^-11^ | 0.73 (0.68~0.78) | 2.59×10^-20^ | 0.75 (0.71~0.81) | 1.32×10^-16^ |
| High | 0.69 (0.64~0.74) | 1.03×10^-26^ | 0.64 (0.60~0.69) | 7.46×10^-37^ | 0.66 (0.61~0.70) | 8.31×10^-33^ |
| *P* for trend | 9.17×10^-26^ | | 1.64×10^-33^ | | 1.44×10^-30^ | |

Note: PA: physical activity; CKD: chronic kidney disease; HR: hazard ratio; CI: confidence interval;

Model 1: crude.

Model 2: adjusted for age, sex, and ethnicity.

Model 3: adjusted for age, sex, ethnicity, smoking status, drinking status, TDI, education, income, and healthy diet score.

Table S9. Association of PA with CKD when excluding participants who developed CKD during the two years of follow-up.

| PA | Model 1 | | Model 2 | | Model 3 | |
| --- | --- | --- | --- | --- | --- | --- |
|  | HR (95% CI) | *P* | HR (95% CI) | *P* | HR (95% CI) | *P* |
| Low | Reference | | Reference | | Reference | |
| Moderate | 0.75 (0.71~0.79) | 2.98×10^-29^ | 0.69 (0.66~0.73) | 4.88×10^-47^ | 0.72 (0.68~0.76) | 5.11×10^-38^ |
| High | 0.66 (0.63~0.70) | 1.04×10^-55^ | 0.62 (0.59~0.65) | 9.93×10^-77^ | 0.64 (0.60~0.67) | 1.72×10^-66^ |
| *P* for trend | 2.01×10^-51^ | | 5.63×10^-68^ | | 3.30×10^-60^ | |

Note: PA: physical activity; CKD: chronic kidney disease; HR: hazard ratio; CI: confidence interval;

Model 1: crude.

Model 2: adjusted for age, sex, and ethnicity.

Model 3: adjusted for age, sex, ethnicity, smoking status, drinking status, TDI, education, income, and healthy diet score.

Table S10. Association of PA with CKD when excluding participants with hypertension and diabetes at baseline.

| PA | Model 1 | | Model 2 | | Model 3 | |
| --- | --- | --- | --- | --- | --- | --- |
|  | HR (95% CI) | *P* | HR (95% CI) | *P* | HR (95% CI) | *P* |
| Low | Reference | | Reference | | Reference | |
| Moderate | 0.82 (0.77~0.87) | 9.16×10^-11^ | 0.75 (0.71~0.80) | 2.65×10^-21^ | 0.77 (0.73~0.82) | 4.13×10^-17^ |
| High | 0.73 (0.68~0.77) | 6.57×10^-25^ | 0.67 (0.63~0.71) | 7.32×10^-39^ | 0.68 (0.64~0.73) | 5.94×10^-34^ |
| *P* for trend | 5.20×10^-20^ | | 3.50×10^-35^ | | 1.56×10^-31^ | |

Note: PA: physical activity; CKD: chronic kidney disease; HR: hazard ratio; CI: confidence interval;

Model 1: crude.

Model 2: adjusted for age, sex, and ethnicity.

Model 3: adjusted for age, sex, ethnicity, smoking status, drinking status, TDI, education, income, and healthy diet score.

Table S11. Association of PA with CKD when further adjusting for the clinical history of hypertension, diabetes, and obesity.

| PA | HR (95% CI) | *P* |
| --- | --- | --- |
| Low | Reference | |
| Moderate | 0.84(0.80~0.87) | 3.19×10^-16^ |
| High | 0.77 (0.74~0.81) | 6.25×10^-29^ |
| *P* for trend | 3.68×10^-27^ | |

Note: PA: physical activity; CKD: chronic kidney disease; HR: hazard ratio; CI: confidence interval. The model was adjusted for age, sex, ethnicity, smoking status, drinking status, TDI, education, income, healthy diet score, and the clinical history of hypertension, diabetes, and obesity.

Table S12. Subgroup analysis for mediation effect of inflammation stratified by age.

| Subgroup | Mediators | *c'* ± *se* | *a* ± *se* | *b* ± *se* | Mediation proportion (95% CI) |
| --- | --- | --- | --- | --- | --- |
| <60 | CRP | -0.111 ± 0.022 | -0.052 ± 0.002 | 0.150 ± 0.012 | 0.06 (0.03~0.08) |
|  | WBC | -0.128 ± 0.022 | -0.038 ± 0.002 | 0.063 ± 0.008 | 0.03 (0.02~0.04) |
|  | INFLA score | -0.108 ± 0.022 | -0.058 ± 0.002 | 0.290 ± 0.021 | 0.11 (0.07~0.16) |
|  | MHR | -0.124 ± 0.022 | -0.035 ± 0.003 | 0.026 ± 0.003 | 0.01 (0.01~0.02) |
| ≥60 | CRP | -0.129 ± 0.013 | -0.053 ± 0.003 | 0.103 ± 0.007 | 0.04 (0.03~0.05) |
|  | WBC | -0.133 ± 0.013 | -0.041 ± 0.003 | 0.069 ± 0.004 | 0.02 (0.01~0.03) |
|  | INFLA score | -0.115 ± 0.013 | -0.055 ± 0.003 | 0.242 ± 0.012 | 0.09 (0.07~0.12) |
|  | MHR | -0.126 ± 0.013 | -0.036 ± 0.002 | 0.136 ± 0.006 | 0.03 (0.02~0.04) |

Note: CRP: C-reactive protein; WBC: white blood cell; INFLA score: a low-grade inflammation; MHR: monocyte to high-density lipoprotein cholesterol ratio; *se*: standard error; CI: confidence interval; *c′* is the effect of PA on CKD after controlling for the impact of inflammation; *a* is the effect of PA on inflammation; *b* is the effect of inflammation on CKD. The mediation model was adjusted for age, sex, ethnicity, smoking status, drinking status, TDI, education, income, and healthy diet score.

Table S13. Subgroup analysis for mediation effect of inflammation stratified by sex.

| Subgroup | Mediators | *c'* ± *se* | *a* ± *se* | *b* ± *se* | Mediation proportion (95% CI) |
| --- | --- | --- | --- | --- | --- |
| Female | CRP | -0.112 ± 0.017 | -0.070 ± 0.003 | 0.136 ± 0.008 | 0.07 (0.04~0.09) |
|  | WBC | -0.129 ± 0.017 | -0.043 ± 0.003 | 0.050 ± 0.004 | 0.02 (0.02~0.03) |
|  | INFLA score | -0.103 ± 0.018 | -0.079 ± 0.003 | 0.277 ± 0.015 | 0.16 (0.11~0.21) |
|  | MHR | -0.125 ± 0.017 | -0.030 ± 0.003 | 0.027 ± 0.003 | 0.01 (0.01~0.02) |
| Male | CRP | -0.128 ± 0.014 | -0.040 ± 0.002 | 0.093 ± 0.008 | 0.03 (0.02~0.04) |
|  | WBC | -0.131 ± 0.014 | -0.035 ± 0.002 | 0.060 ± 0.005 | 0.02 (0.01~0.02) |
|  | INFLA score | -0.116 ± 0.014 | -0.041 ± 0.002 | 0.237 ± 0.014 | 0.07 (0.05~0.09) |
|  | MHR | -0.119 ± 0.014 | -0.041 ± 0.001 | 0.130 ± 0.014 | 0.03 (0.02~0.04) |

Note: CRP: C-reactive protein; WBC: white blood cell; INFLA score: a low-grade inflammation; MHR: monocyte to high-density lipoprotein cholesterol ratio; *se*: standard error; CI: confidence interval; *c′* is the effect of PA on CKD after controlling for the impact of inflammation; *a* is the effect of PA on inflammation; *b* is the effect of inflammation on CKD. The mediation model was adjusted for age, sex, ethnicity, smoking status, drinking status, TDI, education, income, and healthy diet score.

Table S14. Mediation analyses of inflammation within the participants with complete covariate data.

| Mediators | *c'* ± *se* | *a* ± *se* | *b* ± *se* | Mediation proportion (95% CI) | Mediation proportion in the major analysis (95% CI) |
| --- | --- | --- | --- | --- | --- |
| CRP | -0.137 ± 0.014 | -0.049 ± 0.002 | 0.121 ± 0.007 | 0.04 (0.03~0.05) | 0.04 (0.03~0.05) |
| WBC | -0.151 ± 0.014 | -0.036 ± 0.002 | 0.060 ± 0.004 | 0.02 (0.02~0.03) | 0.01 (0.01~0.02) |
| INFLA score | -0.129 ± 0.015 | -0.055 ± 0.002 | 0.282 ± 0.013 | 0.10 (0.08~0.13) | 0.10 (0.08~0.12) |
| MHR | -0.150 ± 0.014 | -0.032 ± 0.002 | 0.031 ± 0.003 | 0.01 (0.01~0.01) | 0.01 (0.01~0.02) |

Note: CRP: C-reactive protein; WBC: white blood cell; INFLA score: a low-grade inflammation; MHR: monocyte to high-density lipoprotein cholesterol ratio; *se*: standard error; CI: confidence interval; *c′* is the effect of PA on CKD after controlling for the impact of inflammation; *a* is the effect of PA on inflammation; *b* is the effect of inflammation on CKD. The mediation model was adjusted for age, sex, ethnicity, smoking status, drinking status, TDI, education, income, and healthy diet score.

Table S15. Mediation analyses of inflammation when excluding participants who developed CKD during the two years of follow-up.

| Mediators | *c'* ± *se* | *a* ± *se* | *b* ± *se* | Mediation proportion (95% CI) | Mediation proportion in the major analysis (95% CI) |
| --- | --- | --- | --- | --- | --- |
| CRP | -0.116 ± 0.011 | -0.048 ± 0.002 | 0.110 ± 0.006 | 0.05 (0.04~0.07) | 0.04 (0.03~0.05) |
| WBC | -0.126 ± 0.011 | -0.035 ± 0.002 | 0.043 ± 0.002 | 0.02 (0.02~0.03) | 0.01 (0.01~0.02) |
| INFLA score | -0.105 ± 0.011 | -0.052 ± 0.002 | 0.252 ± 0.010 | 0.11 (0.08~0.14) | 0.10 (0.08~0.12) |
| MHR | -0.127 ± 0.011 | -0.033 ± 0.002 | 0.030 ± 0.002 | 0.02 (0.01~0.02) | 0.01 (0.01~0.02) |

Note: CRP: C-reactive protein; WBC: white blood cell; INFLA score: a low-grade inflammation; MHR: monocyte to high-density lipoprotein cholesterol ratio; *se*: standard error; CI: confidence interval; *c′* is the effect of PA on CKD after controlling for the impact of inflammation; *a* is the effect of PA on inflammation; *b* is the effect of inflammation on CKD. The mediation model was adjusted for age, sex, ethnicity, smoking status, drinking status, TDI, education, income, and healthy diet score.

Table S16. Mediation analyses of inflammation when excluding participants with hypertension and diabetes at baseline.

| Mediators | *c'* ± *se* | *a* ± *se* | *b* ± *se* | Mediation proportion (95% CI) | Mediation proportion in the major analysis (95% CI) |
| --- | --- | --- | --- | --- | --- |
| CRP | -0.096 ± 0.013 | -0.051 ± 0.002 | 0.114 ± 0.007 | 0.04 (0.02~0.05) | 0.04 (0.03~0.05) |
| WBC | -0.105 ± 0.013 | -0.038 ± 0.002 | 0.051 ± 0.004 | 0.02 (0.01~0.02) | 0.01 (0.01~0.02) |
| INFLA score | -0.089 ± 0.013 | -0.056 ± 0.002 | 0.234 ± 0.012 | 0.10 (0.08~0.13) | 0.10 (0.08~0.12) |
| MHR | -0.105 ± 0.013 | -0.036 ± 0.002 | 0.030 ± 0.002 | 0.02 (0.01~0.02) | 0.01 (0.01~0.02) |

Note: CRP: C-reactive protein; WBC: white blood cell; INFLA score: a low-grade inflammation; MHR: monocyte to high-density lipoprotein cholesterol ratio; *se*: standard error; CI: confidence interval; *c′* is the effect of PA on CKD after controlling for the impact of inflammation; *a* is the effect of PA on inflammation; *b* is the effect of inflammation on CKD. The mediation model was adjusted for age, sex, ethnicity, smoking status, drinking status, TDI, education, income, and healthy diet score.

Table S17. Mediation analyses of inflammation when further adjusting for the clinical history of hypertension, diabetes, and obesity.

| Mediators | *c'* ± *se* | *a* ± *se* | *b* ± *se* | Mediation proportion (95% CI) | Mediation proportion in the major analysis (95% CI) |
| --- | --- | --- | --- | --- | --- |
| CRP | -0.072 ± 0.011 | -0.037 ± 0.001 | 0.094 ± 0.006 | 0.04 (0.03~0.06) | 0.04 (0.03~0.05) |
| WBC | -0.079 ± 0.011 | -0.028 ± 0.002 | 0.040 ± 0.003 | 0.02 (0.01~0.02) | 0.01 (0.01~0.02) |
| INFLA score | -0.069 ± 0.011 | -0.040 ± 0.004 | 0.198 ± 0.010 | 0.10 (0.07~0.14) | 0.10 (0.08~0.12) |
| MHR | -0.113 ± 0.013 | -0.036 ± 0.002 | 0.028 ± 0.002 | 0.01 (0.01~0.02) | 0.01 (0.01~0.02) |

Note: CRP: C-reactive protein; WBC: white blood cell; INFLA score: a low-grade inflammation; MHR: monocyte to high-deok

nsity lipoprotein cholesterol ratio; *se*: standard error; CI: confidence interval; *c′* is the effect of PA on CKD after controlling for the impact of inflammation; *a* is the effect of PA on inflammation; *b* is the effect of inflammation on CKD. The mediation model was adjusted for age, sex, ethnicity, smoking status, drinking status, TDI, education, income, healthy diet score, and the clinical history of hypertension, diabetes, and obesity.

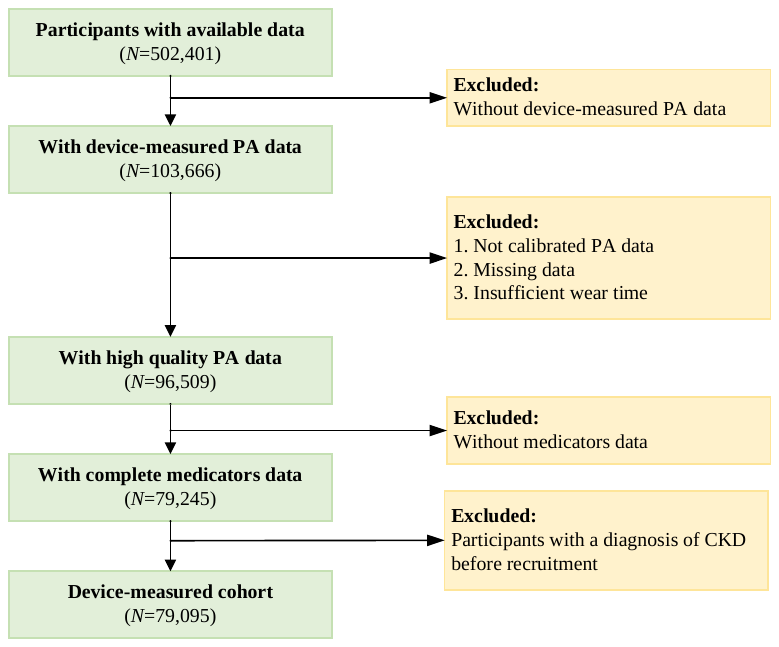

Figure S1. Specific selection process for participants in the device-measured cohort.

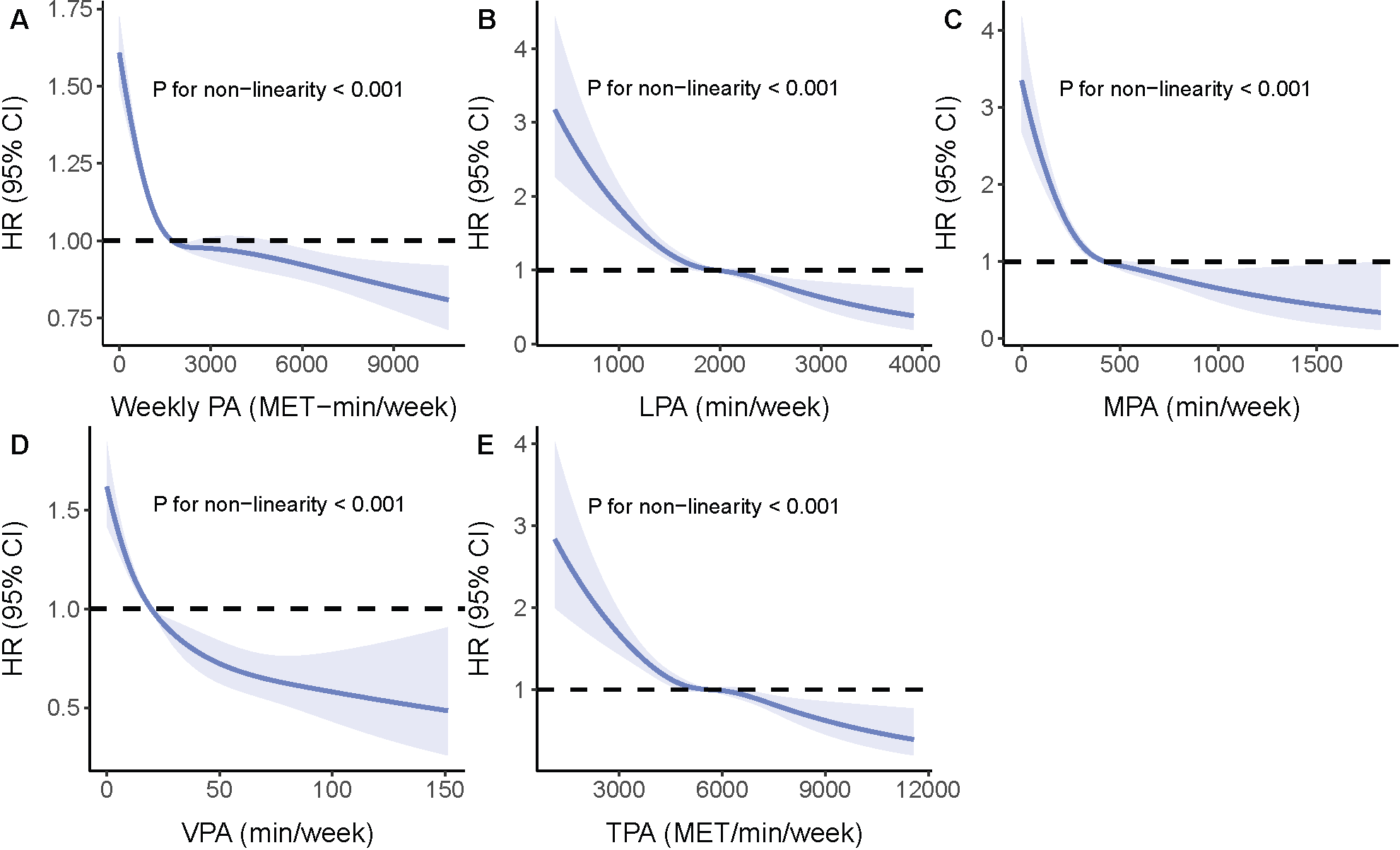

Figure S2. Restricted cubic spline curves for the influence of PA on CKD in the two different UK Biobank cohorts.

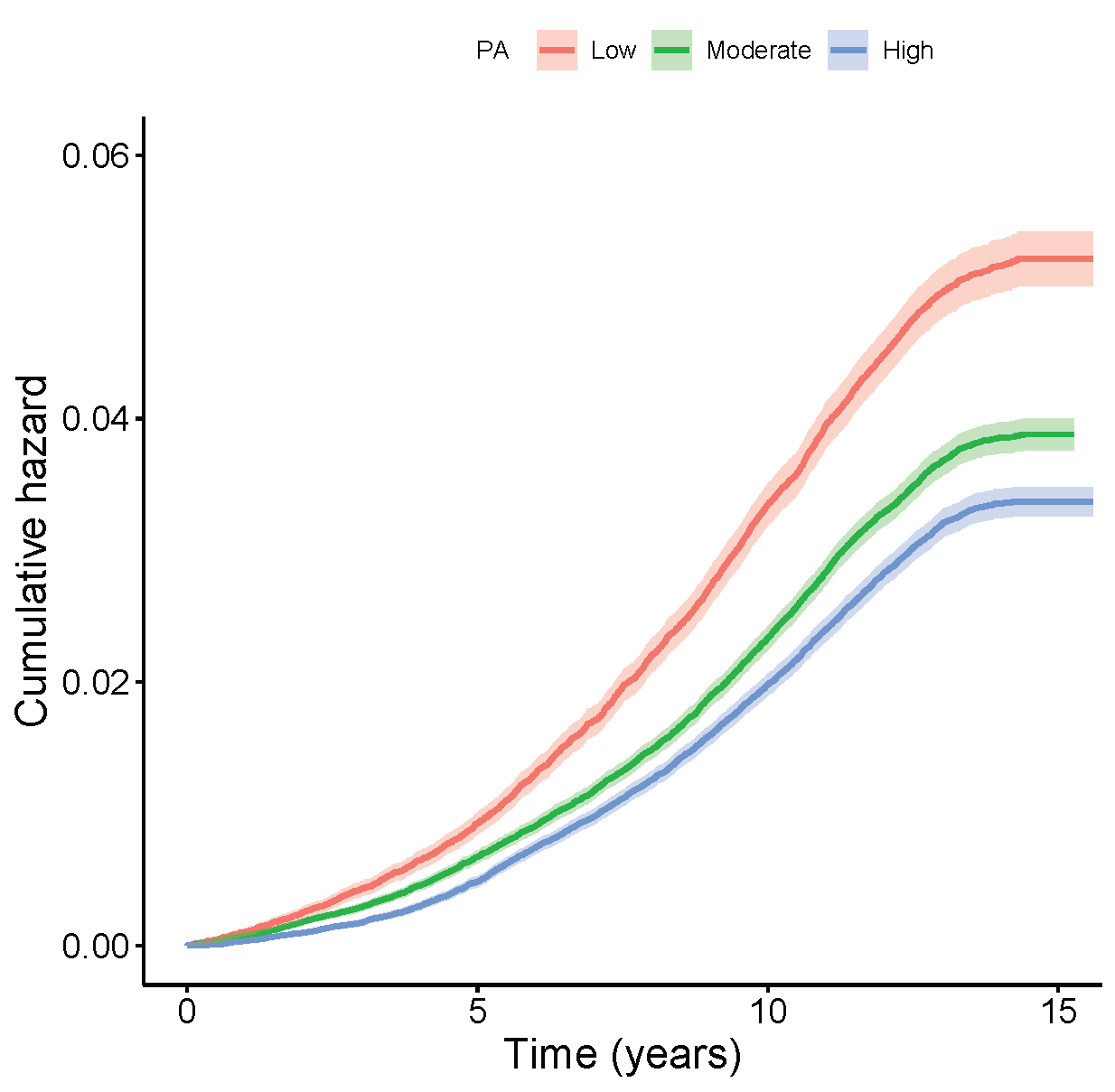

Figure S3. Kaplan-Meier curves for the influence of different levels of PA on CKD in the questionnaire-measured cohort.
